# Interventions designed to improve vaccination uptake: Scoping review of systematic reviews and meta-analyses - protocol (version 1)

**DOI:** 10.1101/2021.08.18.21262232

**Authors:** CJ Heneghan, A Plüddemann, EA Spencer, J Brassey, EC Rosca, IJ Onakpoya, DH Evans, JM Conly, NT Brewer, T Jefferson

**Affiliations:** The University of Oxford, Radcliffe Observatory Quarter, Oxford, OX2 6GG, UK; Trip Database Ltd. Glasllwch Lane, Newport, NP20 3PS, UK; Victor Babes University of Medicine and Pharmacy of Timisoara Pia□a Eftimie Murgu 2, Timi□oara 300041, Romania; Li Ka Shing Institute of Virology and Dept. of Medical Microbiology & Immunology, University of Alberta; Departments of Medicine, Microbiology, Immunology & Infectious Diseases, Pathology & Laboratory Medicine, Synder Institute for Chronic Diseases and O’Brien Institute for Public Health, Cumming School of Medicine, University of Calgary and Alberta Health Services, Calgary, Canada; Gillings School of Global Public Health and Lineberger Comprehensive Cancer Center. University of North Carolina

**Keywords:** Vaccine uptake, Vaccine hesitancy, Immunization, systematic reviews

## Abstract

**Background:** Vaccine uptake varies substantially, and resources to promote the uptake of vaccines differ widely by country and income level. As a result, immunization rates are often suboptimal. There is a need to understand what works, particularly in low- and middle-income countries and other settings where resources are scarce.

**Methods:** We plan to conduct a scoping review of interventions designed to increase vaccination uptake

We will include systematic reviews and meta-analyses of interventional studies that address the question of vaccine uptake. We will search the following electronic databases: MEDLINE, Cochrane Database of Systematic Reviews, EMBASE, Epistemonikos, Google Scholar, LILACs and TRIP database (which covers guidelines and the grey literature) until 01 July 2021 and hand-search the reference lists of included articles. We will include systematic reviews that comprise studies of all ages if they report quantitative data on the impact on vaccine uptake. To assess the quality, we will use a modified AMSTAR score and ate the quality of the evidence in included reviews using the “Grade of Recommendations Assessment, Development and Evaluation” (GRADE).

**Expected results:** We intend to present the evidence using summary tables to present the evidence stratified by vaccine coverage, the specific population, e.g., children, adolescents and older adults, and by setting, e.g. healthcare, community. We will also present when low middle-income subgroups are reported.

## Background

Vaccine uptake varies substantially by age, gender, ethnicity, geographical location, and socioeconomic status. Research has established that some of these differences are due to variations in vaccination’s behavioural and social drivers (BeSD). [1] In addition, the resources required to successfully promote the uptake of vaccines varies widely by country and income level. As a result, immunization programmes often struggle to achieve optimal coverage of the target population.

WHO seeks to complement its ongoing work on the measurement of BeSD with information on effective interventions to increase vaccine uptake. This information can help vaccination programmes to understand what works for whom and in what settings, particularly in low- and middle-income countries and other locations where resources are scarce.

Therefore, we plan to do a scoping review of systematic reviews of published evidence on interventions designed to increase vaccine uptake. We will categorize the interventions and their components across different populations and geographical regions. The WHO plans to use the scoping review results to guide immunization programmes’ selection of interventions to promote vaccine uptake.

## Methods

### Objectives

To conduct a scoping review of interventions designed to increase vaccination uptake.

#### The specific focus of the review

- Population: e.g., children, adolescents, adults and older adults (age 65+;
- Interventions: any intervention designed to improve participation in vaccination;
- Outcomes: Uptake, Hesitancy, Disease risk appraisal, Confidence, Social norms, Provider recommendation, Availability.

#### Type of studies

We will include systematic reviews and meta-analyses of interventional studies that address the question of vaccine uptake.

#### Search strategy

We will search the following electronic databases: MEDLINE, Cochrane Database of Systematic Reviews, EMBASE, Epistemonikos, Google Scholar, LILACs and TRIP database (which covers guidelines and the grey literature) until 01 July 2021 and hand-search the reference lists of included articles. The searches will combine free and thesaurus search terms and keywords related to vaccine uptake (vaccin* OR innocul OR immunis* OR immuniz*). In the first instance, we will use sensitive search filters developed by the Health Information Research Unit at McMaster University, Canada, to focus on systematic reviews and meta-analyses. [2] We will also search the bibliographies of retrieved systematic reviews. We will screen all titles and abstracts of retrieved citations for inclusion. Based on the results of the initial filter for systematic reviews, we will review the need for further search terms (see Appendix for sample search terms). The final search strategy will be developed with advice from information specialists and an iterative process adapted for each database. Two reviewers will independently evaluate the full text of articles potentially meeting eligibility criteria. Discrepancies will be resolved through discussion. Where a consensus cannot be reached, a third reviewer will arbitrate.

#### Eligibility criteria

We will include systematic reviews that comprise studies of all ages if they report quantitative data on the impact on vaccine uptake. In addition, we will include systematic reviews that contain randomized controlled trials (RCTs) and quasi-experimental (including interrupted time series and before-and-after studies). We will exclude reviews that assess only vaccine efficacy or effectiveness and reviews that do not include any RCTs.

#### Types of interventions

Interventions that aim to increase vaccine uptake in a specific population or the overall population.

#### Quality assessment

To assess the quality, we will use modified AMSTAR score items 3 and 7. [3] Item 3: “Was a comprehensive* literature search performed?” At least two electronic sources should be searched. The report must include years and databases used (e.g. Central, EMBASE, and MEDLINE), plus keywords or MESH terms. Item 7: “Was the scientific quality of the included studies assessed and documented?” ‘A priori’ assessment methods should be provided (e.g., for effectiveness studies if the author(s) chose to include only randomized, double-blind, placebo-controlled studies, or allocation concealment as inclusion* consistent with a systematic review search.

One reviewer will record and assess the reporting of the quality of included systematic reviews and report the assessments, and a second reviewer will independently check the quality ratings. Disagreements will be resolved through discussion with a third reviewer, who will arbitrate where a consensus cannot be reached. We will rate the quality of the evidence in included reviews using the “Grade of Recommendations Assessment, Development and Evaluation” (GRADE). [4] We will downgrade or upgrade the rating for the quality of the evidence, based on the amount of potential bias due to study design and other criteria specified in the GRADE, and provide a summary of findings tables by the outcomes of interest. GRADE assessment will be based on assessing the risk of bias and an evaluation of inconsistency, indirectness, and imprecision of the results and other factors (see the GRADE Table in the Appendix for more information). We will check the rating, where GRADE has been used to assess primary studies included in the reviews. Where another tool has been used to assess quality, one reviewer will convert this to a GRADE assessment, and a second reviewer will independently check the assessment.

#### Data extraction

We will conduct the review according to PRISMA (Preferred Reporting Items for Systematic Reviews and Meta-Analyses) guidelines. [5] Data from included reviews will be extracted by one reviewer and independently checked by a second reviewer. We will structure the outcomes by intervention type and create categories using an iterative process to extract objectives and self-reported outcomes. In addition, we will extract data on the population, study characteristics (e.g., number of trials, location etc.) and the intervention and comparator and the outcomes of interest as well as the type of meta-analysis effect model used in the meta-analysis (fixed or random) and between-study heterogeneity estimates (I^2^ values).

Where two reviews cover the same intervention and outcome with overlapping studies, we will select the most relevant review (i.e. more comprehensive and up-to-date) for inclusion; and include the historical reviews in an appendix. We will also use the Jadad decision algorithm to interpret discordant reviews and select the most appropriate review evidence for interventions (see figure 1). Two authors will independently apply the algorithm to reach a consensus over which review/meta-analysis is included. [6]

**Fig. 1:**
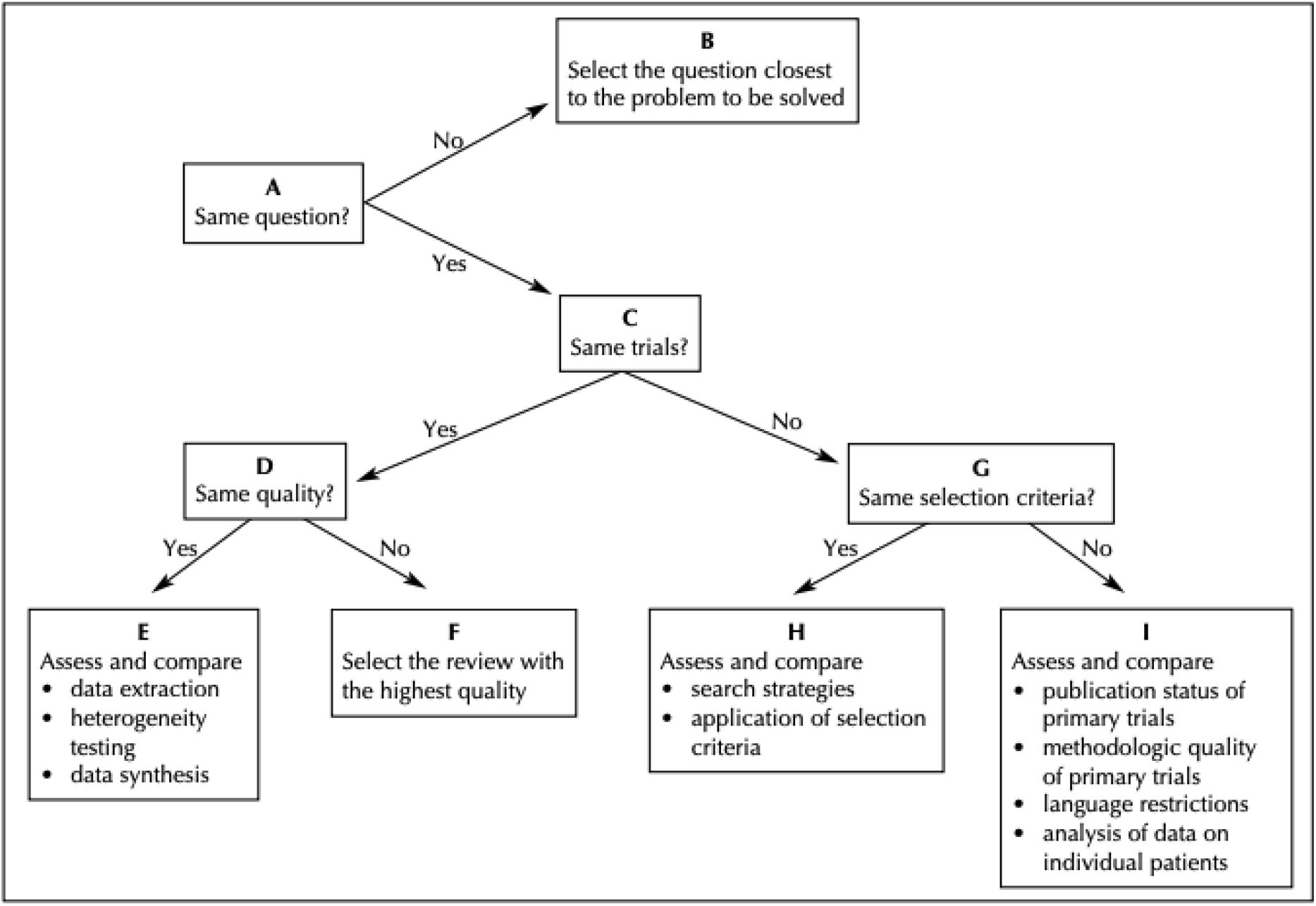
A decision algorithm for interpreting discordant reviews (assuming that the reviews have few and minimal flaws).

#### Outcomes of interest

We will prioritize outcomes according to the WHO handbook for guideline development [7] as High (critical for decision making), Moderate (important for decision making) and Low (not important for decision making). Vaccination uptake is the highest priority. Constructs in the WHO BeSD Framework are deemed moderate. Other constructs not in the framework are low priority.

**Table 1.** Outcomes of interest and prioritization.

| Priority | Construct | Definition | Includes |
| --- | --- | --- | --- |
| High | Uptake | Receipt of vaccine | Initiation, completion, coverage and behaviour |
| Moderate | Hesitancy | Motivational state of being conflicted about, or opposed to, getting vaccinated | Intentions, willingness, openness, stage |
| Moderate | Disease risk appraisal | The belief that one is at risk for an infectious disease that vaccination can prevent | Perceived risk, perceived susceptibility, concern, perceived severity, anticipated regret, worry, fear |
| Moderate | Confidence | Attitudes and beliefs that vaccines work are safe and are part of a trustworthy medical system | Perceived importance, benefit, and effectiveness; concerns about safety, harm, side effects, and adverse events; trust in providers and the system of vaccination |
| Moderate | Social norms | Shared expectations of acceptable vaccination behaviour by a group | Descriptive norms, injunctive norms |
| Moderate | Provider recommendation | Advice from a health care worker to receive vaccination | Advice in clinical and non-clinical settings |
| Moderate | Availability | Low/unavailable stocks may play a role in vaccine uptake | The vaccine is unavailable in the country or just the specific clinic |
| Moderate | Access | Perceived and actual access to immunization services | Distance, travel, timing, location, ease, convenience, opportunity costs, financial costs |
| Low | Knowledge | An accurate understanding of facts about vaccination | It does not include awareness, but we will assess it where reported |

We will use summary tables to present the evidence stratified by vaccine coverage, the specific population, e.g., children, adolescents and older adults, and by setting, e.g. healthcare, community. We will also present when low middle-income subgroups are reported. We will present the data as reported in the paper and. Where significant heterogeneity exists, assessed using expert judgement, we will extract the reasons. Once the data is mapped out, we will attempt to present the findings for categories of interventions mapped onto the BeSD Framework. The Thinking and Feeling domain will include interventions that use education, confidence building, persuasion, motivational interviewing, and decision aids. The Social Processes domain will include interventions that use social norms, social networks, altruism and healthcare provider communication. The Practical Factors domain will include interventions that use reminder/recall, implementation intentions, mere measurement, default appointments, onsite vaccination (including work and school), incentives, requirements (mandates), and sanctions.

## Data Availability

All data in the manuscript are made available

https://figshare.com/s/5416371b9164af1ed716

## Authors’ contributions

All authors contributed in equal part to the conceptualization and development of the content. TJ and CH wrote the first draft and edited this version. All authors contributed to the subsequent drafts and approved the final version. The authors acknowledge the substantial intellectual contribution of Dr Lisa Menning, Dr Julie Leask, and the WHO BeSD working group in conceptualizing the research question, outcomes, and intervention types.

## Conflict of interest statements

TJ received a Cochrane Methods Innovations Fund grant to develop guidance on the use of regulatory data in Cochrane reviews (2015 to 2018). From 2014 to 2016, he was a member of three advisory boards for Boehringer Ingelheim. TJ was a member of an independent data monitoring committee for a Sanofi Pasteur clinical trial on an influenza vaccine. Market research companies occasionally interview TJ about phase I or II pharmaceutical products for which he receives fees (current). TJ was a member of three advisory boards for Boehringer Ingelheim (2014 to 16). TJ was a member of an independent data monitoring committee for a Sanofi Pasteur clinical trial on an influenza vaccine (2015 to 2017). TJ is a relator in a False Claims Act lawsuit on behalf of the United States that involves sales of Tamiflu for pandemic stockpiling. If resolved in the United States favour, he would be entitled to a percentage of the recovery. TJ is coholder of a Laura and John Arnold Foundation grant for the development of a RIAT support centre (2017 to 2020) and Jean Monnet Network Grant, 2017 to 2020 for The Jean Monnet Health Law and Policy Network. TJ is an unpaid collaborator to the Beyond Transparency in Pharmaceutical Research and Regulation led by Dalhousie University and funded by the Canadian Institutes of Health Research (2018 to 2022). TJ consulted for Illumina LLC on next-generation gene sequencing (2019 to 2020). TJ was the consultant scientific coordinator for the HTA Medical Technology programme of the Agenzia per I Servizi Sanitari Nazionali (AGENAS) of the Italian MoH (2007 to 2019). TJ is Director Medical Affairs for BC Solutions, a market access company for medical devices in Europe. TJ was funded by NIHR UK and the World Health Organization (WHO) to update Cochrane review A122, Physical Interventions to interrupt the spread of respiratory viruses. Oxford University funds TJ to carry out a living review on the transmission epidemiology of COVID 19. Since 2020, TJ receives fees for articles published by The Spectator and other media outlets. TJ is part of a review group carrying out a Living rapid literature review on the modes of transmission of SARS CoV 2 (WHO Registration 2020/1077093 0). He is a member of the WHO COVID 19 Infection Prevention and Control Research Working Group, for which he receives no funds. TJ is funded to co-author rapid reviews on the impact of Covid restrictions by the Collateral Global Organisation.

CJH holds grant funding from the NIHR, the NIHR School of Primary Care Research, the NIHR BRC Oxford and the World Health Organization for a series of Living rapid reviews on the modes of transmission of SARs CoV 2 reference WHO registration No2020/1077093. He has received financial remuneration from an asbestos case and given legal advice on mesh and hormone pregnancy tests cases. He has received expenses and fees for his media work, including occasional payments from BBC Radio 4 Inside Health and The Spectator. He receives expenses for teaching EBM and is also paid for his GP work in NHS out of hours (contract Oxford Health NHS Foundation Trust). He has also received income from the publication of a series of toolkit books and appraising treatment recommendations in non-NHS settings. He is the Director of CEBM, an NIHR Senior Investigator and an advisor to Collateral Global.

DE holds grant funding from the Canadian Institutes for Health Research and Li Ka Shing Institute of Virology relating to the development of Covid 19 vaccines and the Canadian Natural Science and Engineering Research Council concerning Covid 19 aerosol transmission. He is a recipient of World Health Organization and Province of Alberta funding which supports the provision of BSL3 based SARS CoV 2 culture services to regional investigators. He also holds public and private sector contract funding relating to the development of poxvirus based Covid 19 vaccines, SARS CoV 2 inactivation technologies, and serum neutralization testing.

JMC holds grants from the Canadian Institutes for Health Research on acute and primary care preparedness for COVID 19 in Alberta, Canada and was the primary local Investigator for a Staphylococcus aureus vaccine study funded by Pfizer, for which all funding was provided only to the University of Calgary. He is a co-investigator on a WHO funded study using integrated human factors and ethnography approaches to identify and scale innovative IPC guidance implementation supports in primary care with a focus on low resource settings and using drone aerial systems to deliver medical supplies and PPE to remote First Nations communities during the COVID 19 pandemic. He also received support from the Centers for Disease Control and Prevention (CDC) to attend an Infection Control Think Tank Meeting. He is a member of the WHO Infection Prevention and Control Research and Development Expert Group for COVID 19 and the WHO Health Emergencies Programme (WHE) Ad hoc COVID 19 IPC Guidance Development Group, both of which provide multidisciplinary advice to the WHO, for which no funding is received and from which no funding recommendations are made for any WHO contracts or grants. He is also a member of the Cochrane Acute Respiratory Infections Group.

JB is a major shareholder in the Trip Database search engine (www.tripdatabase.com) as well as being an employee. In relation to this work, Trip has worked with a large number of organizations over the years; none have any links with this work. The main current projects are with AXA and Collateral Global.

ECR was a member of the European Federation of Neurological Societies(EFNS) / European Academy of Neurology (EAN) Scientist Panel, Subcommittee of Infectious Diseases (2013 to 2017). Since 2021, she is a member of the International Parkinson and Movement Disorder Society (MDS) Multiple System Atrophy Study Group, the Mild Cognitive Impairment in Parkinson Disease Study Group, and the Infection Related Movement Disorders Study Group. She was an External Expert and sometimes Rapporteur for COST proposals (2013, 2016, 2017, 2018, 2019) for Neurology projects. She is a Scientific Officer for the Romanian National Council for Scientific Research.

NB serves as a paid consultant for the US Centers for Disease Control and Prevention, World Health Organization, and Merck. He is a member of the Lancet Commission on Vaccine Acceptance in the United States. He is a member of the COVID-19 Vaccine Communications Advisory Group, North Carolina Department of Health and Human Services. He is a Member of the Vaccine Confidence Work Group, Board of Scientific Counselors, Deputy Director of Infectious Disease, Centers for Disease Control and Prevention, 2020-present. He is a member of the Vaccine Confidence Working Group, National Vaccine Advisory Committee. He is a member of the Research Working Group, Vaccine Acceptance Research Network.

IJO, EAS, and AP have no interests to disclose.

## Ethics committee approval

No ethics approval was necessary.

## Data Availability

All data included in the review will be provided in the tables and text.

## Appendix

### Search terms

Additional search terms that we might include but not be limited to are: (vaccine OR innocul OR immunis* OR immuniz*) ^s1^

AND

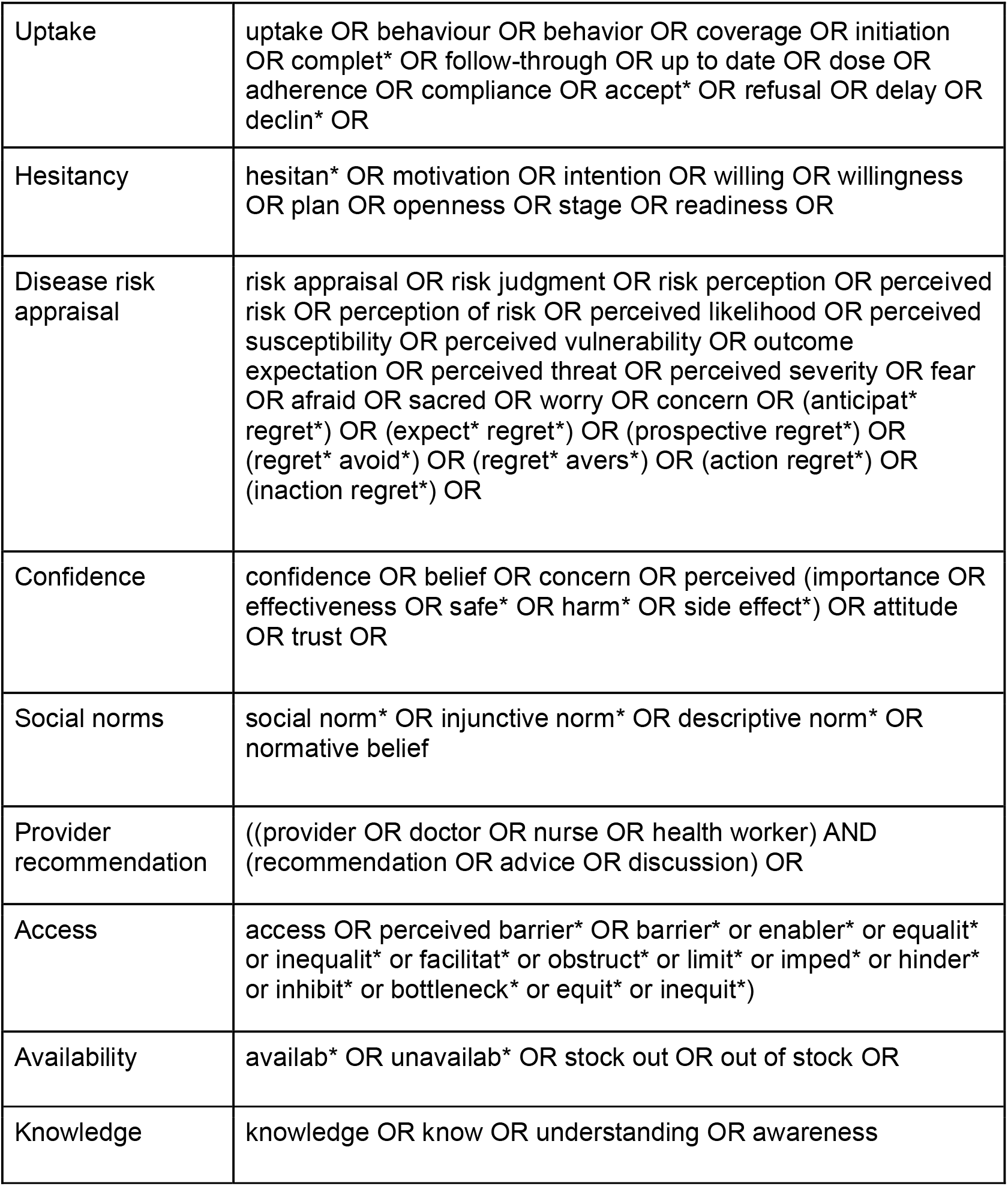

The research team considered including a third grouping of search terms for intervention (e.g., intervention* OR reminder* OR incentive*). However, we prefer to have a larger body of studies and not to narrow the search results in this way.

S1: ‘Vaccination coverage’ is also a Medical Subject Heading (MeSH) term in Medlin GRADE tables.

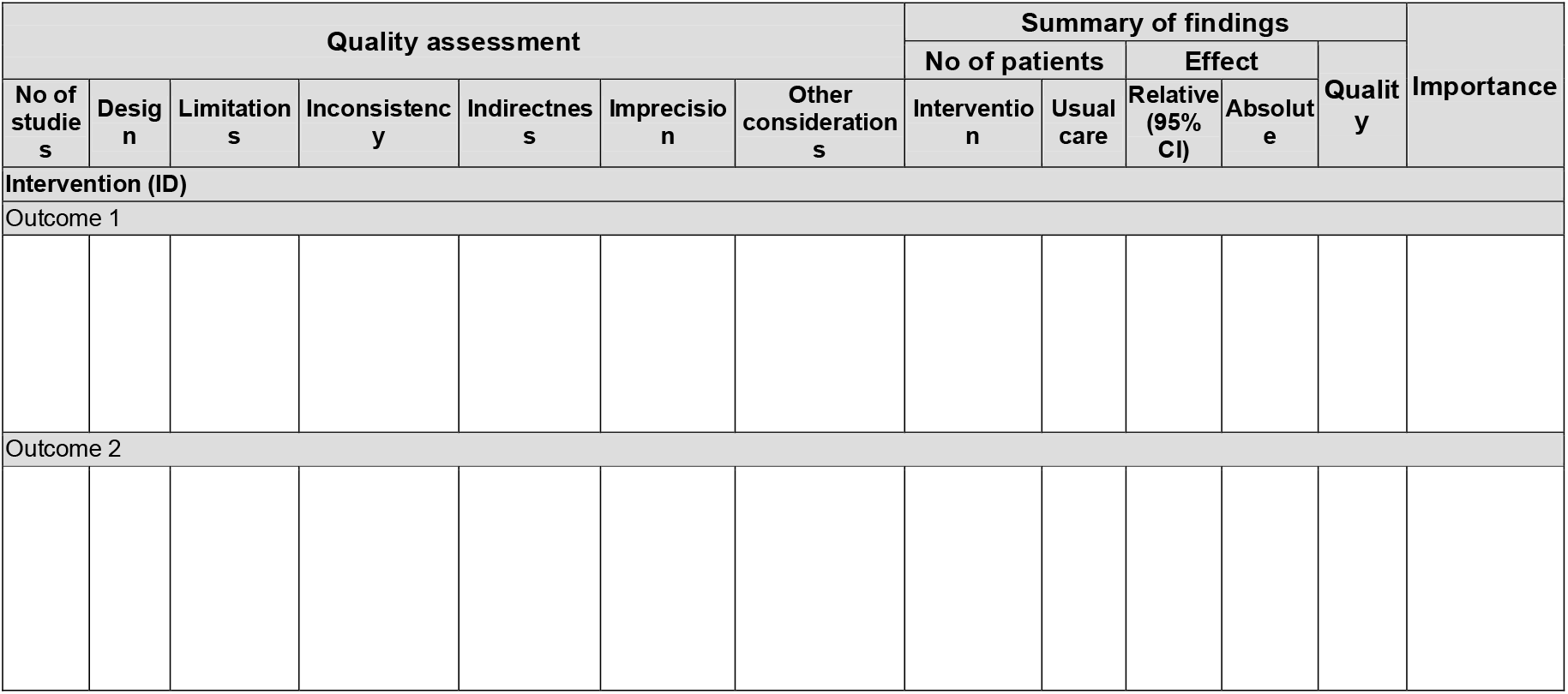

- **Limitations - assessing risk of bias**
  ○ Lack of allocation concealment: Those enrolling patients are aware of the group (or period in a crossover trial) to which the next enrolled patient will be allocated (major problem in “pseudo” or “quasi” randomized trials with allocation by day of week, birth date, chart number, etc)
  ○ Lack of blinding: Patient, care givers, those recording outcomes, those adjudicating outcomes, or data analysts are aware of the arm to which patients are allocated (or the medication currently being received in a crossover trial)
  ○ Incomplete accounting of patients and outcome events: Loss to follow-up and failure to adhere to the intention-to-treat principle in superiority trials; or in non-inferiority trials, loss to follow-up, and failure to conduct both analyses considering only those who adhered to treatment, and all patients for whom outcome data are available
  ○ Selective outcome reporting bias: Incomplete or absent reporting of some outcomes and not others on the basis of the results
  ○ Other limitations: Stopping early for benefit; Use of unvalidated outcome measures (e.g., patient-reported outcomes); Carryover effects in crossover trial; Recruitment bias in cluster-randomized trials
- **Inconsistency**
  ○ Reviewers should consider rating down for inconsistency when: 1.Point estimates vary widely across studies; 2.Confidence intervals (CIs) show minimal or no overlap;3.The statistical test for heterogeneity—which tests the null hypothesis that all studies in a meta-analysis have the same underlying magnitude of effect—shows a low P-value; 4.The I2—which quantifies the proportion of the variation in point estimates due to among-study differences—is large.
- **Indirectness**
  ○ We are more confident in the results when we have direct evidence. By direct evidence, we mean research that directly compares the interventions in which we are interested delivered to the populations in which we are interested and measures the outcomes important to patients. Thus, we can have concerns about indirectness when the population, intervention, or outcomes differ from those in which we are interested. In general, evidence based on surrogate outcomes should usually trigger rating down, whereas the other types of indirectness will require a more considered judgment.
- **Imprecision**
  ○ When considering the quality of evidence, the issue is whether the CI around the estimate of treatment effect is sufficiently narrow. If it is not, we rate down the evidence quality by one level (for instance, from high to moderate). If the CI is very wide, we might rate down by two levels.
- **Other**
  ○ A number of factors may rate the quality of evidence up or down. These include presence or absence of publication bias, when a large magnitude of effect exists, when there is a dose–response gradient, and when all plausible confounders or other biases increase our confidence in the estimated effect.

